# On temporal changes in the role of different age groups in propagating the Omicron epidemic waves in England

**DOI:** 10.1101/2022.12.30.22283949

**Authors:** Edward Goldstein

## Abstract

**Background:** There is limited information on the role of individuals in different age groups in the spread of infection during the Omicron epidemics, especially ones beyond the winter epidemic wave in 2021-2022. COVID-19 booster vaccination in England during the Autumn 2022 was restricted to persons aged over 50y, and persons in clinical risk groups.

**Methods:** We used previously developed methodology to evaluate the role of individuals in different age groups in propagating the Spring, Summer, and Autumn waves of the Omicron epidemic in England. This methodology utilizes the relative risk (RR) statistic that measures the change in the proportion of cases in each age group among all COVID-19 cases in the population before the peak of an epidemic wave vs. after the peak of an epidemic wave. Higher values for the RR statistic represent age groups that experienced a disproportionate depletion of susceptible individuals during the ascent of the epidemic (due to increased contact rates and/or susceptibility to infection).

**Results:** For the 2022 Spring wave, the highest RR estimate belonged to children aged 5-9y (RR=2.05 (95%CI (2.02,2.08)), followed by children aged 10-14y (RR=1.68 (1.66,1.7)) and children aged 0-4y (RR=1.38 (1.36,1.41)). For the Summer wave, the highest RR estimates belonged to persons aged 20-34y: (RR=1.09 (1.07,1.12) in aged 20-24y, RR=1.09 (1.07,1.11) in aged 25-29y, RR=1.09(1.07,1.11) in aged 30-34y). For the Autumn wave, the highest RR estimates belonged to those aged 70-74y (RR=1.10 (1.07,1.14)), followed by adults aged 35-39y (RR=1.09 (1.06,1.12)), adults aged 40-44y (RR=1.09 (1.06,1.12)), and adults aged 65-69y (RR=1.08 (1.05,1.11)).

**Conclusions:** As time progressed, the greatest relative roles in propagating different waves of the Omicron epidemic in England shifted from school-age children to younger adults to adults aged 35-44y and 65-74y. Extending booster vaccination to all adults, and possibly to children should help limit the spread of Omicron infections in the community.

## Introduction

The proliferation of the Omicron variant of SARS-CoV-2 resulted in the decline in the rates of severe outcomes, including hospitalizations and deaths associated with SARS-CoV-2 infections. The risk of those outcomes in infected adults became lower compared to the Delta variant, though relative risks for severe outcomes for Omicron vs. Delta vary with age, being highest for the oldest adults [1,2]. Additionally, the relative contribution of different age groups to the transmission of SARS-CoV-2 infection in the community changed with the appearance of the Omicron variant. For prior SARS-CoV-2 variants, the leading role in the acquisition and transmission of infection generally belonged to younger adults (aged 18-35y), as well as older adolescents [3-6]. The emergence of the Omicron variant saw an increase in the rates of infection in children compared to other age groups. For example, for respiratory samples tested between January 5-20, 2022 in England, the greatest prevalence of infection belonged children aged 5-11y [7], which wasn’t the case for previous SARS-CoV-2 variants [6]. The modeling study for the SARS-CoV-2 epidemic during the winter of 2021-2022 in England also estimated that the highest incidence rates around the peak of the epidemic belonged to children [8], with adults playing a more prominent role during the early stages of that epidemic wave in December of 2021. Less is known about the role of different age groups during the subsequent waves of Omicron epidemics, in England and elsewhere. Coronavirus (COVID-19) Infection Surveys in the UK (e.g. [9-11]) provide data on the percentage of people testing positive for coronavirus (COVID-19) in private residential households. However, studies of SARS-CoV-2 viral shedding suggest that viral shedding is quite prolonged, and its duration is positively correlated with age [12-13]. Therefore, percentage of people testing positive for coronavirus [9-11] overestimates the rate of incidence of recent infection in different age groups, particularly for older persons. Our study of the Spring, Summer and Autumn Omicron epidemics in France suggests the importance of children and younger adults in the spread of Omicron infections [14]. However, the relative role of children in the spread of Omicron infections in France declined during the Autumn wave of the 2022 epidemic compared to the earlier waves [14], and the relative role of children might be even smaller in places that experienced higher rates of infection in children earlier on.

The burden of mortality with a SARS-CoV-2 infection during the Spring, Summer and Autumn epidemic waves in England remained relatively high compared to the 2021-2022 Winter wave [15], which supports the utility of vaccination and other mitigation measures for limiting the spread of Omicron infection in the community. In particular, there is evidence that vaccines both reduce the risk of Omicron infection [16], as well as the risk of onward transmission of infection [17]. Following the emergence of the Omicron variant, booster vaccinations for adults, as well as for children were offered in England [18,19], though for the autumn of 2022 the booster vaccination program was restricted to persons aged over 50y, as well as persons in clinical risk groups [20]. Very recently, the booster vaccination program was extended to persons aged 18-49y [21].

In our earlier work [22-24] we introduced a method for evaluating the role of individuals in different population groups in the spread of infection and applied it to data from epidemics associated with influenza, respiratory syncytial virus (RSV) and pertussis in the United States. That method calculates, for each age group, the relative risk (RR) statistic that reflects the change in the proportion of cases in that age group among all detected cases of disease in the population for the period before the epidemic peak vs. the period after the epidemic peak. That method is used to characterize age groups that experienced a disproportionate depletion of susceptible individuals during the ascent period of the epidemic (due to increased contact rates and/or susceptibility to infection). Moreover, simulations for influenza epidemics [22] suggested that age groups with higher values for the RR statistics were generally also the age groups for which vaccination yielded the greatest effect on reducing the epidemic’s growth rate. We applied this method to data from SARS-CoV-2 epidemics in 2020 and 2021 [3,4] to show that younger adults and older adolescents had the greatest relative role in the spread of infection in the corresponding locations. We also applied this method to the Spring, Summer and Fall waves of the Omicron epidemic in France [14] and found that the greatest relative role in the spread of infection belonged to children aged 10-19y, particularly during periods when schools were open, followed by children aged 0-9y and adults aged 20-29y, as well as adults aged 30-49y. In this paper, apply the method in [3,4,22-24] to Omicron epidemics in England to better understand the role of individuals in different age groups in the spread of infection during the subsequent waves of Omicron epidemics beyond the initial (winter) wave.

## Methods

### Data

Data on the daily number of COVID-19 cases in different age groups in England are available from [15].

### Statistical Inference

Data on detected COVID-19 cases in England ([15], and Figure 1 in this paper) suggest three epidemic waves between Feb. 15, 2022 -- Dec. 15, 2022: The Spring wave (with a peak of March 21 for the number of detected COVID-19 cases), the Summer wave (with a peak of July 4), and the Autumn wave (with the peak of Oct. 3). For each epidemic wave, we excluded the 7-day period around the peak day for detected COVID-19 cases from the analyses. We defined the before-the-peak period for an epidemic wave as the 14-day period prior to the 7-day window around the peak day for COVID-19 cases, and the after-the-peak period of the epidemic wave as the 14-day period starting from the first day after the 7-day window around the peak of COVID-19 cases. Thus, for the Spring epidemic wave with the peak of March 21, the excluded period around the peak is March 18-24, the before-the-peak period used in our study is March 4 – March 17, and the after-the-peak period for that wave is March 25 – April 7, etc.

**Figure 1:**
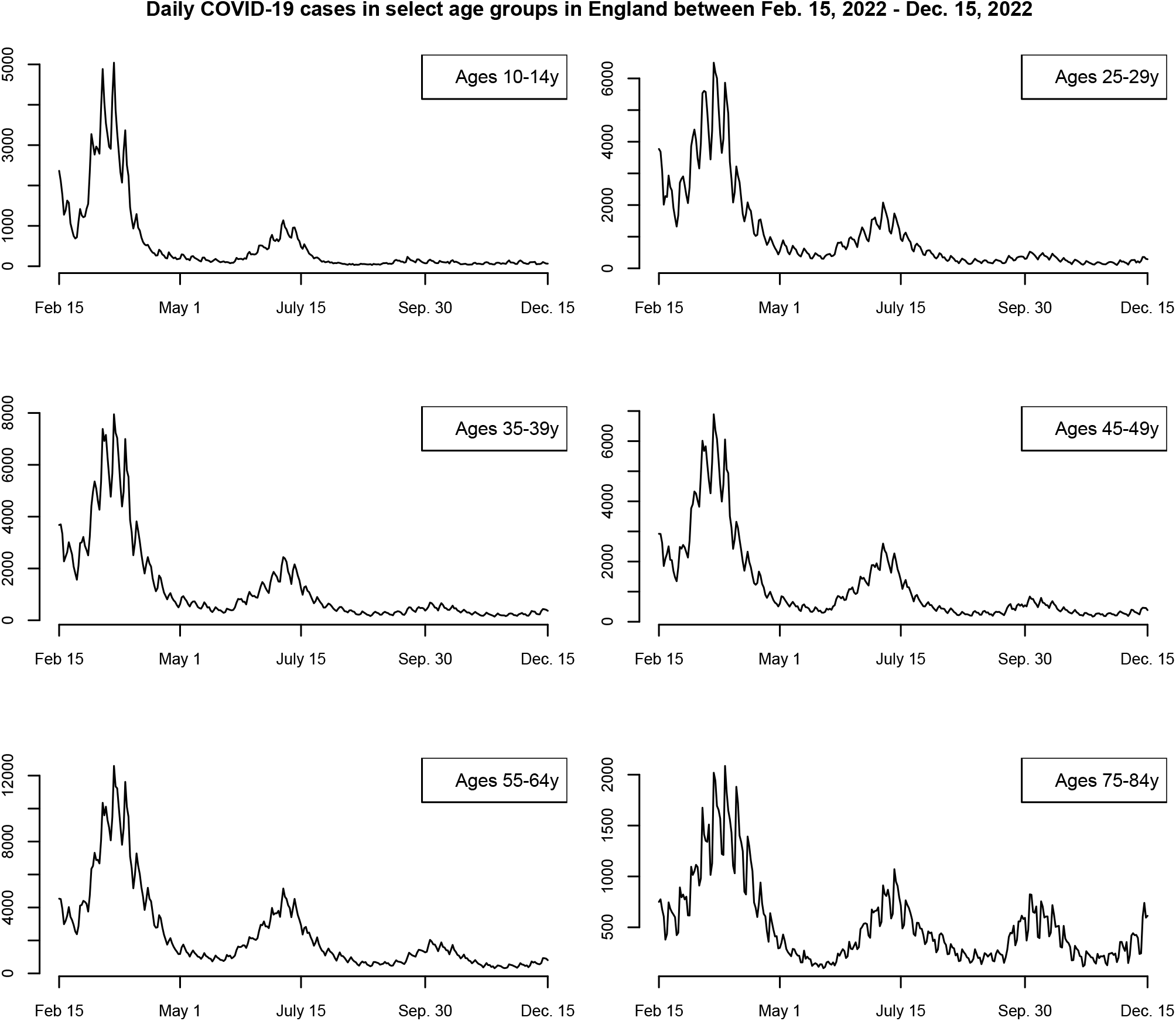
Daily COVID-cases in select age groups in England between Feb. 15, 2022 - Dec. 15, 2022.

For the Spring and the Summer epidemic waves, we considered 15 age groups in our analyses: 0-4y, 5-9y, 10-14y, 15-19y, 20-24y, 25-29y, 30-34y, 35-39y, 40-44y, 45-49y, 50-55y, 55-64y, 65-74y, 75-84y, 85+y. Figure 1 suggests that case detection was limited/irregular during the Autumn wave for younger age groups. For the Autumn epidemic wave, we considered 14 age groups of adults: 25-29y, 30-34y, 35-39y, 40-44y, 45-49y, 50-54y, 55-59y, 60-64y, 65-69y, 70-74y, 75-79y, 80-84y, 85-89y, 90+y. While we’ve excluded children from our analyses for the Autumn wave of the epidemic, infection rates in children in England during that period were lower compared to adults [11], with children likely not playing the leading roles in propagating that epidemic wave. For each epidemic wave, and each age group *g*, let *B*(*g*) represent the number of COVID-19 cases in individuals in the age group *g* during the before-the-peak-period, and let *A*(*g*) represent the number of COVID-19 cases in individuals in the age group *g* during the after-the-peak-period. The proportion of cases in the age group *g* among all cases during the before-the-peak period is therefore

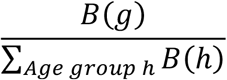

with the corresponding estimate for the after-the-peak period. The relative risk (RR) is:

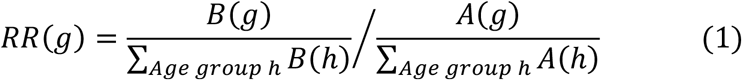

Higher value for *RR*(*g*) suggests a decline in the proportion of cases in the age group *g* during the after-the-peak period due to a disproportionate depletion of susceptible individuals in the age group *g* during the ascent period of the epidemic (due to increased contact rates and/or increased susceptibility to infection). As the numbers of reported cases in different age groups in England are sufficiently high [15], the logarithm ln(*RR*(*g*)) of the relative risk *RR*(*g*) is approximately normally distributed [24], with the standard error *SE*(*g*) for ln(*RR*(*g*)) being

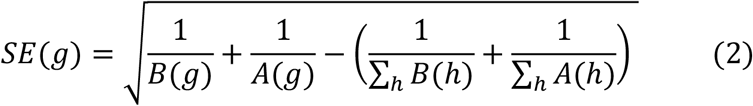

## Results

Figure 1 plots the daily numbers of detected COVID-19 cases in England in select age groups between Feb. 15, 2022 - Dec. 15, 2022, suggesting three epidemic waves during our study period. Figure 1 suggests that as time progressed, a smaller proportion of COVID-19 cases got detected, particularly in younger age groups.

Table 1 gives the estimates of the relative risk (RR) statistic in different age groups during the Spring and the Summer epidemic waves, while Table 2 gives the estimates of the relative risk (RR) statistic in different age groups of adults during the Autumn wave of the Omicron epidemic in England. For the Spring wave, the highest RR estimate belonged to children aged 5-9y (RR=2.05 (95%CI (2.02,2.08)), followed by children aged 10-14y (RR=1.68 (1.66,1.7)) and children aged 0-4y (RR=1.38 (1.36,1.41)). For the Summer wave, the highest RR estimates belonged to persons aged 20-34y (RR=1.09 (1.07,1.12) in aged 20-24y, RR=1.09 (1.07,1.11) in aged 25-29y, RR=1.09 (1.07,1.11) in aged 30-34y). For the Autumn wave, the highest RR estimate in adults belonged to those aged 70-74y (RR=1.10 (1.07,1.14)), followed by adults aged 35-39y (RR=1.09 (1.06,1.12)), adults aged 40-44y (RR=1.09 (1.06,1.12)), and adults aged 65-69y (RR=1.08 (1.05,1.11)).

**Table 1:** Estimates for the relative risk RR (eq. 1) for data on COVID-19 cases in different age groups during the Spring and Summer waves of the Omicron epidemic in England in 2022.

| Age group | RR for the Spring Wave | RR for the Summer Wave |
| --- | --- | --- |
| 0-4y | 1.38 (1.36,1.41) | 0.84 (0.8,0.88) |
| 5-9y | 2.05 (2.02,2.08) | 1 (0.95,1.04) |
| 10-14y | 1.68 (1.66,1.7) | 0.99 (0.97,1.02) |
| 15-19y | 1.21 (1.19,1.23) | 1.07 (1.04,1.1) |
| 20-24y | 1.01 (1,1.02) | 1.09 (1.07,1.12) |
| 25-29y | 1.05 (1.04,1.06) | 1.09 (1.07,1.11) |
| 30-34y | 1.06 (1.05,1.07) | 1.09 (1.07,1.11) |
| 35-39y | 1.1 (1.09,1.11) | 1.02 (1,1.04) |
| 40-44y | 1.11 (1.1,1.12) | 1.04 (1.02,1.05) |
| 45-49y | 1.01 (1,1.02) | 1.05 (1.03,1.07) |
| 50-54y | 0.9 (0.9,0.91) | 1.04 (1.03,1.06) |
| 55-64y | 0.83 (0.83,0.84) | 1.02 (1.01,1.03) |
| 65-74y | 0.77 (0.76,0.78) | 0.98 (0.97,0.99) |
| 75-84y | 0.69 (0.68,0.7) | 0.78 (0.76,0.79) |
| 85+y | 0.63 (0.61,0.64) | 0.67 (0.66,0.69) |

**Table 2:** Estimates for the relative risk RR (eq. 1) for data on COVID-19 cases in different age groups of adults during the Autumn wave of the Omicron epidemic in England in 2022.

| Age group | Relative Risk (RR) |  | Age group | Relative Risk (RR) |
| --- | --- | --- | --- | --- |
| 25-29y | 1.07 (1.03,1.1) |  | 60-64y | 0.95 (0.93,0.98) |
| 30-34y | 1.03 (1,1.07) |  | 65-69y | 1.08 (1.05,1.11) |
| 35-39y | 1.09 (1.06,1.12) |  | 70-74y | 1.10 (1.07,1.14) |
| 40-44y | 1.09 (1.06,1.12) |  | 75-79y | 0.98 (0.95,1.01) |
| 45-49y | 1.01 (0.98,1.03) |  | 80-84y | 0.9 (0.86,0.93) |
| 50-54y | 0.96 (0.94,0.99) |  | 85-89y | 0.92 (0.89,0.96) |
| 55-59y | 0.93 (0.91,0.96) |  | 90+y | 0.83 (0.8,0.87) |

## Discussion

The appearance of the Omicron variant of SARS-CoV-2 resulted both in changes in the severity of infections [1,2], as well as in changes in the contribution of different age groups to the transmission of SARS-CoV-2 infection in the population. For the earlier SARS-CoV-2 variants, the leading role in the spread of infection generally belonged to younger adults (aged 20-35y), as well as older adolescents [3-6]. For the 2021-2022 winter wave of the Omicron epidemic in England, infection rates in children were higher relative to other age groups compared to earlier variants (e.g. the results of the REACT-1 studies [7] vs. [6]). Less is known about the contribution of different age groups to the spread of infection during the subsequent waves of the Omicron epidemic. In this paper, we used the previously developed methodology [3,4,22-24] to evaluate the role of individuals in different age groups in the spread of infection during the Spring, Summer, and Autumn waves of the Omicron epidemic in England. For the Spring epidemic wave, we found a disproportionate depletion of susceptible individuals during the ascent period of the epidemic in children aged 5-14y, followed by children aged 0-4y and 15-19y. This result is generally in agreement with the findings in our study of Omicron epidemics in France [14], as well as with the high rates/earlier peaks of infection in children in England in the UK coronavirus survey data for that period [9]. We also note that several studies ([26-28]) suggest significant spread of SARS-CoV-2 infection in the school setting under limited mitigation of transmission (which generally applies to the Omicron period compared to the circulation of the earlier SARS-CoV-2 variants). The role of children in the spread of infection declined during the summer (presumably due to immunity acquired during the previous epidemic waves, as well as the fact that schools were closed), which is in agreement with the data on infections for England in the UK coronavirus survey data for that period [10]. The greatest relative role in propagating the Summer Omicron epidemic in England belonged to adults aged 20-34y. The greatest relative role in propagating the Autumn Omicron epidemic in England belonged to adults aged 65-74y and 35-44y.

The SARS-CoV-2-associated mortality burden during the spring, summer and autumn epidemics in England remained significant compared to the mortality burden during the winter wave of the Omicron epidemic [15], with many deaths with a detected SARS-CoV-2 infection in England during the Omicron period not having COVID-19 on the death certificate (compare the different figures in [29]). Additionally, there is evidence about a substantial burden of Omicron-associated deaths for which SARS-CoV-2 infection wasn’t detected [30]. This, together with evidence that vaccines both reduce the risk of Omicron infection [16], as well as the risk of onward transmission of infection [17], suggests the utility of including all adults, and possibly children, in the booster vaccination program. We note that booster COVID-19 vaccination during the Autumn of 2022 in England was offered only to persons aged over 50y and persons in clinical risk groups [19], though subsequently it was extended to adults aged 18-49y [21].

Our results have some limitations. Temporal changes in the detection of Omicron infections during the course of an epidemic wave would affect the estimates of the RR statistic in different age groups. We chose 5-week periods around the peak of each epidemic wave as our study period to minimize the effect of changes in case-detection during the course of each epidemic wave, with shorter periods around the epidemic peak yielding similar estimates for the relative role of different age groups. There is uncertainty regarding the interpretation of the RR statistic in terms of the role that individuals in different age groups play in the spread of infection.

Simulations for influenza epidemics showed that higher values for the RR statistic generally correspond to age groups for which vaccination has a bigger impact on reducing the epidemic’s growth rate [22].

## Conclusions

We found that as time progressed, the greatest relative roles in propagating different waves of the Omicron epidemic in England shifted from school-age children to younger adults to adults aged 35-44y and 65-74y. Extending the booster vaccination program in England to adults aged under 50y, and possibly to children should help limit the spread of Omicron infections in the community and mitigate the Omicron-associated mortality burden, which remains substantial [29].

## Data Availability

This study is based on aggregate, de-identified publicly available data that can be retrieved through ref. 15.

https://coronavirus.data.gov.uk/

